# Diagnostic yield of tongue swab- compared to sputum-based molecular testing for tuberculosis in four high-burden countries

**DOI:** 10.1101/2025.07.03.25330836

**Authors:** Caitlin A. Moe, Rita Kabuleta Luswata, Armen Jheannie Barrameda, Hien Le, Seke Muzazu, Rebecca Crowder, Alfred O. Andama, Claudia M. Denkinger, Monde Muyoyeta, Ha Phan, Adithya Cattamanchi, Charles Yu, the TSwaY Study Network

## Abstract

**Background:** Tongue swabs are a promising alternative specimen for tuberculosis (TB) diagnosis, with high specificity (>98%) but lower sensitivity than sputum-based molecular tests. We investigated whether increased sample availability could offset lower sensitivity, resulting in similar diagnostic yield.

**Methods:** From September 2024-January 2025, we screened consecutive people with presumptive TB at health centers in the Philippines, Vietnam, Uganda, and Zambia. Enrolled participants were asked to provide tongue swabs and referred for routine sputum collection. Tongue swabs were tested in research laboratories using the MiniDock MTB Test (Guangzhou Pluslife Biotech Co., Ltd., China); sputum was tested using WHO-recommended molecular testing per national guidelines. We compared diagnostic yield, defined as positive test results among those seeking testing, between tongue swab- and sputum-based molecular testing with a pre-specified non-inferiority margin of ±3%.

**Findings:** Of 1,639 participants, 851 (51·9%) were female, 415 (25·3%) were living with HIV and 132 (8·1%) were children <5 years. Overall, 1,389 (84·7%) provided sputum, while 1,637 (99·9%) provided tongue swabs. Diagnostic yield was similar for tongue swab-(63/1639, 3·8%) and sputum-based (68/1639, 4·1%) molecular testing, and within the pre-specified non-inferiority margin (difference -0·3%, 95%CI -1·2 to +0·6). Results were consistent across subgroups by country, age, sex, and HIV status.

**Interpretation:** TB diagnostic yield from tongue swabs was non-inferior to sputum-based molecular testing. These data support scale-up of swab-based molecular platforms as a lower-cost alternative to sputum-based tests, particularly in settings where sputum collection is challenging, or smear microscopy remains standard.

**Funding:** Supported by the Bill and Melinda Gates Foundation.

**RESEARCH IN CONTEXT:** *Evidence before this study:*

- Tongue swabs are a promising alternative specimen type for tuberculosis (TB) diagnosis.
- Studies have demonstrated high specificity (>98%) but lower sensitivity on tongue swabs than sputum-based molecular tests.
- Recent studies highlight that less sensitive non-sputum tests could have similar or even higher diagnostic yield due to the increased likelihood of people offered testing being able to provide a sample.
- No previous studies have directly compared the TB diagnostic yield of tongue swabs to sputum.

*Added value of this study:*

- This is the first comparative study of the diagnostic yield of tongue swab- vs. sputum-based molecular testing, conducted in real-world clinical settings across four high TB burden countries.
- Using a low-cost molecular platform, we found that tongue swabs achieved similar diagnostic yield to sputum, with differences within a pre-specified non-inferiority margin.
- Notably, tongue swabs were successfully collected from nearly all participants, including children and people living with HIV—groups in whom sputum collection can be especially difficult.

*Implications of all the available evidence:*

- TB diagnostic yield from tongue swabs is non-inferior to sputum-based molecular testing, demonstrating the value of tongue swabs as a viable and potentially lower-cost alternative to sputum for TB diagnosis.
- These findings support further scale up and operational research to integrate tongue swab testing into TB diagnostic algorithms, particularly in settings where sputum collection is challenging, or smear microscopy remains the primary diagnostic method.

## INTRODUCTION

Rapid diagnosis and treatment of tuberculosis (TB) is critical for reducing morbidity and mortality, as well as minimizing disease transmission. However, TB remains the leading infectious killer worldwide in part because 2·7 million of the 10·8 million people estimated to develop TB annually are not diagnosed and treated.^1^ More than half of TB diagnostic units in high burden countries do not have current World Health Organization (WHO)-recommended rapid diagnostics (WRDs) due to cost and infrastructure requirements.^2^ Further, prevalence studies have revealed that a substantial proportion of people with TB have not yet sought care at health facilities where testing would be available.^3^ Others seek care but are unable to spontaneously produce sputum. To reduce global TB burden, there is an urgent need for lower cost and simpler diagnostics from an accessible sample type that can be deployed in peripheral health centers and enable point-of-care (POC) testing as part of community-based active case finding activities.^4^

Tongue swabs are emerging as a promising alternative specimen type for TB diagnosis. Studies have shown that TB bacilli on the tongue dorsum can be detected using molecular assays.^5^ In contrast to sputum, tongue swabs are easy to collect, including from young children and people with no or minimal symptoms.^6,7^ Moreover, swabs are simpler to process than sputum: Swab-based molecular testing for TB does not require nucleic acid extraction or purification, which add cost and complexity to sputum-based tests.^8^ For example, MiniDock MTB Test (Guangzhou Pluslife Biotech Co., Ltd., China) provides results in 12-25 minutes using a portable, battery-operated platform that is anticipated to be substantially less expensive than current sputum based WRDs.^9,10^

Swab-based platforms are expected to expand the reach of molecular testing for TB to lower-level health facilities. Preliminary studies, however, suggest that sensitivity for TB detection using tongue swabs may be lower compared to sputum-based molecular testing.^6,11,12^ Steadman et al. found that tongue swab MiniDock MTB was more sensitive than sputum smear microscopy (85·7% vs. 67·1%, p=0.001) but less sensitive than sputum Xpert Ultra (80·6% vs. 92·8%, p<0.001).^9^ Despite this, recent studies highlight that less sensitive non-sputum tests could have similar or even higher diagnostic yield due to the increased likelihood of people offered testing being able to provide a sample.^13^ To date, there are no comparative studies of the diagnostic yield of tongue swab- vs. sputum-based molecular testing.

To support policy and uptake of tongue swab-based molecular tests, we conducted a multi-country study among people presenting to outpatient health centers. Our primary objective was to compare the diagnostic yield of tongue swab-based testing using the MiniDock MTB Test to routine sputum-based molecular testing using WRDs. Secondarily, we assessed earlier steps in the testing cascade (e.g., sample provision) and preferences related to specimen collection for TB testing.

## METHODS

### Study Design and Participants

In this pragmatic cross-sectional study at outpatient health centers in the Philippines, Vietnam, Uganda, and Zambia, we screened consecutive people of all ages presenting to study health centers and included those with presumptive TB based on: 1) unexplained cough for 2 or more weeks or 2) a TB risk factor (TB contact, history of mining, or HIV infection) plus an abnormal WHO-endorsed TB screening test (chest x-ray, or for PLHIV, serum C-reactive protein [CRP] >5 mg/dL). We also included children <10 years old with cough for less than 2 weeks plus unexplained fever for ≥2 weeks, unexplained weight loss/poor weight gain, unexplained failure to thrive, severe acute malnutrition, or unexplained lethargy or reduced playfulness for ≥ 2 weeks. We excluded people if they were presenting for TB treatment after being diagnosed with TB elsewhere, already receiving treatment for TB, started on treatment for extra-pulmonary TB only, or unable or unwilling to provide informed consent.

### Study Procedures

Trained research staff collected brief demographic and clinical data (age, sex, HIV status) from each participant (or from a caregiver for young children). Participants were asked to provide a tongue swab and then referred for sputum collection. Tongue swabs (Copan FLOQswab 520CS01) were collected by trained research staff into proprietary tubes pre-filled with buffer provided with the MiniDock MTB Test kit (Guangzhou Pluslife Biotech Co., Ltd., China). Participants were requested to not eat or drink for at least 30 minutes prior to tongue swab collection. Using a back-front and left-right motion, each tongue was swabbed from the back of the top of the tongue and as far back as possible without creating a gag reflex (about ¾ of the visible tongue dorsum) for 30 seconds. Immediately after collection, the tongue swab was inserted into the proprietary pre-filled tube and the swab head was swirled against the bottom and sides of the tube 10 times. The swab head was then pinched by squeezing the outside of the tube and discarded. Tubes were closed and transported to a research laboratory for testing within 24 hours of collection. Sputum collection (or stool collection for younger children) was performed by routine health center staff per local guidelines. After being asked to provide tongue swab and sputum samples, participants completed a brief survey on their experience with and preference for tongue swab vs. sputum collection.

### Molecular testing for TB

Testing of tongue swab samples with MiniDock MTB Test was performed within 24 hours of collection by research laboratory staff in accordance with manufacturer recommendations. Briefly, tubes were placed in the Pluslife Thermolyse device for 5 minutes, the lysed sample was added to the MiniDock MTB Test card and the test card was inserted into the MiniDock device per manufacturer instructions (see **Supplement** for details). After about 25 minutes, a result was displayed with indicator lights on the MiniDock device as either positive, negative, or invalid.

Molecular testing of sputum samples was performed by routine health center staff in accordance with manufacturer recommendations using Xpert MTB/RIF Ultra (Cepheid, USA), or at one health center in Uganda, Truenat MTB Plus (Molbio Diagnostics, India). Sputum results were considered positive if they had a semi-quantitative grade of ‘Very Low’ or higher.

### Outcome Measures

The primary outcome was the diagnostic yield of molecular testing, defined as the proportion of people with a TB-positive test result from a sample collected on the day of enrollment, among those enrolled (*i.e.,* seeking testing for presumptive TB). Secondary outcomes included earlier steps in the TB testing cascade (proportion who provided a sample for molecular testing on the day of enrollment and proportion with a valid molecular test result), the proportion who experienced discomfort when providing a sample, and participant preference for sputum vs. tongue swab collection.

### Sample Size and Statistical Analysis

Using power simulation methods for McNemar’s test of paired proportions, we calculated that a sample size of 1,600 participants (400 per country) would achieve at least 90% power to assess the non-inferiority of tongue swab-based molecular testing compared to sputum-based molecular testing. This calculation assumed a non-inferiority margin of ± 3%, a TB prevalence of 10%, a sensitivity of 75% for tongue swab-based molecular testing, a 25% rate or individuals unable to provide sputum samples, and a two-sided alpha of 0·05.

Following the prespecified analysis plan, we summarized all outcomes for tongue swab vs. sputum samples as proportions with 95% confidence intervals (CI). We compared outcome proportions for tongue swab vs. sputum overall and for key subgroups (country, sex, age strata and HIV status) using McNemar’s test of paired proportions. In a sensitivity analysis, we compared diagnostic yield when classifying “Trace” sputum Xpert Ultra results as positive. Analyses were conducted in Stata version 18 (StataCorp, College Station, TX).

### Ethics Statement

Written informed consent was obtained from all study participants. Ethical approval for the study was obtained from institutional review boards and/or research ethics committees at the University of California San Francisco (USA), University of Heidelberg (Germany), De la Salle Medical and Health Sciences Institute (Philippines), Uganda National Council for Science and Technology (Uganda), National Lung Hospital (Vietnam), and University of Zambia (Zambia).

## RESULTS

### Study Population

Between September 2024 and January 2025, 6244 people were screened for presumptive TB across all study sites and 4605 (73·8%) were excluded, mostly (n=3,995 [86·8%]) because they did not meet symptom or risk factor eligibility criteria (**Figure 1**). Of the 1,639 participants enrolled, 851 (51·9%) were female, 415 (25·3%) were living with HIV and 132 (8·1%) were children <5 years old (**Table 1**).

**Figure 1.**
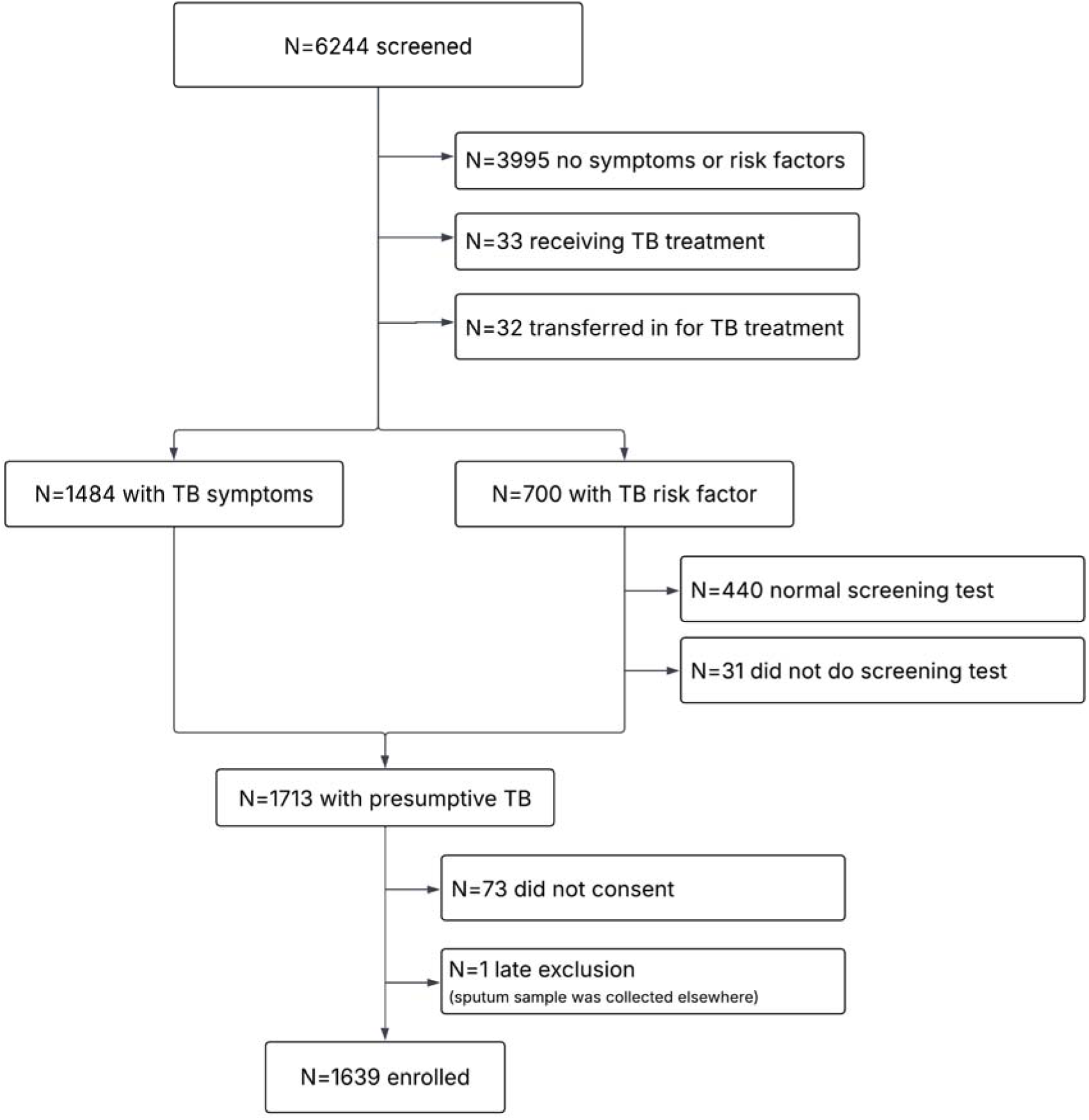
Participant flow diagram.

**Table 1.** Participant characteristics.

|  | **Total**<br>(N=1639) | **Philippines**<br>(N=273) | **Vietnam**<br>(N=650) | **Uganda**<br>(N=300) | **Zambia**<br>(N=416) |
| --- | --- | --- | --- | --- | --- |
| *Age group* |  |  |  |  |  |
| <5 years | 132 (8.1%) | 47 (17.2%) | 8 (1.2%) | 38 (12.7%) | 39 (9.4%) |
| 5-11 years | 107 (6.5%) | 35 (12.8%) | 9 (1.4%) | 33 (11.0%) | 30 (7.2%) |
| 12-18 years | 86 (5.2%) | 30 (11.0%) | 18 (2.8%) | 21 (7.0%) | 17 (4.1%) |
| 19-64 years | 1170 (71.4%) | 137 (50.2%) | 522 (80.3%) | 196 (65.3%) | 315 (75.7%) |
| ≥65 years | 144 (8.8%) | 24 (8.8%) | 93 (14.3%) | 12 (4.0%) | 15 (3.6%) |
| Female sex<sup>a</sup> | 851 (51.9%) | 151 (55.3%) | 315 (48.5%) | 195 (65.0%) | 190 (45.7%) |
| *HIV status* |  |  |  |  |  |
| Positive | 415 (25.3%) | 0 (0.0%) | 190 (29.2%) | 45 (15.0%) | 180 (43.3%) |
| Negative | 913 (55.7%) | 233 (85.3%) | 201 (30.9%) | 255 (85.0%) | 224 (53.8%) |
| Unknown | 311 (19.0%) | 40 (14.7%) | 259 (39.8%) | 0 (0.0%) | 12 (2.9%) |
| History of TB | 237 (14.5%) | 42 (15.4%) | 105 (16.2%) | 8 (2.7%) | 82 (19.7%) |
| Cough ≥ 2 weeks | 1370 (83.6%) | 235 (86.1%) | 475 (73.1%) | 286 (95.3%) | 374 (89.9%) |

<sup>a</sup> missing for n=2 who declined to report

### Diagnostic Yield

Diagnostic yield was similar for tongue swab-(63/1639; 3·8%) and sputum-based (68/1639; 4·1%) molecular testing, and the difference was within the pre-specified margin of non-inferiority (difference -0·3%; 95% CI -1·2 to +0·6) (**Figure 2** and **Table S1**). There was no significant difference in diagnostic yield in sub-groups defined by country, HIV status, sex and age (**Table S1**). The difference in diagnostic yield between tongue swab- vs. sputum-based molecular testing was within the pre-specified 3% margin of non-inferiority in three of four countries, people living with and without HIV, males and females, and most age groups. Of note, the diagnostic yield of combined tongue swab and sputum molecular testing was higher (90/1639; 5·5%) than with either specimen type alone.

**Figure 2.**
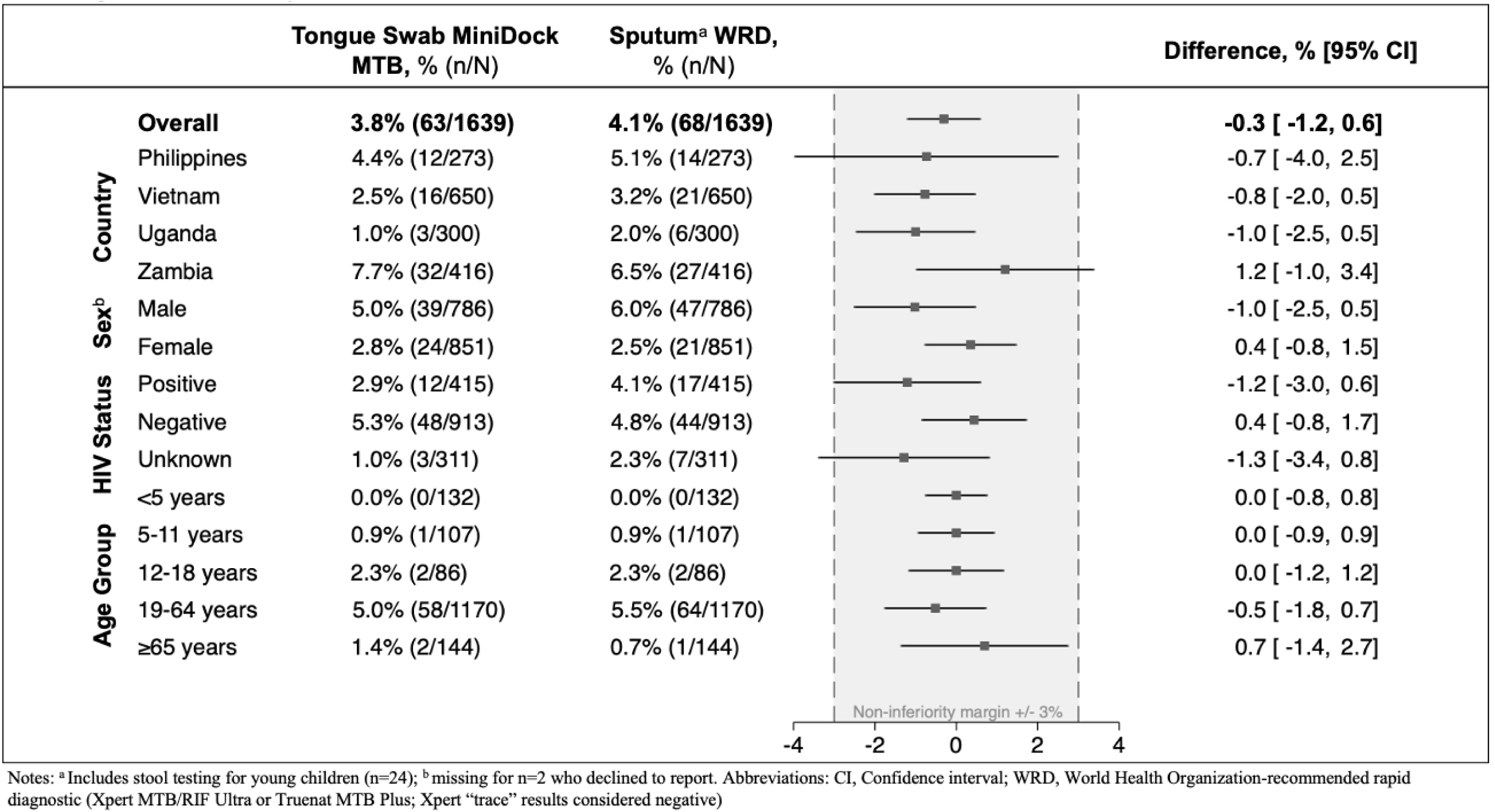
Difference in diagnostic yield of tongue swab- vs. sputum-based molecular testing for TB. The plot shows the difference (black square) in diagnostic yield and its 95% confidence interval (whiskers). The shadow box shows the pre-specified ±3% margin of non-inferiority.

### Secondary Outcomes

Overall, 1,389 (84·7%) provided sputum or stool on the day of presentation (24 children aged 0-7 years provided stool). The proportion of participants who provided same-day tongue swabs (n=1,637/1,639; 99·9%) was higher than the proportion who provided sputum or stool (difference 15·1%; 95% CI 13·3-16·9) (**Table 2**). Results varied by country (difference ranging from 2·0% to 35·5%) but were similar for other subgroups (**Supplemental Tables S2-S5**). Of note, among the 1,389 (84·7%) participants who provided sputum (excluding 24 stool samples), 1,006 (73·7%) provided a salivary sample, 199 (14·6%) provided a mucoid sample, 139 (10·2%) provided a mucopurulent sample, and 17 (1·3%) provided a purulent sample; 4 (0·3%) sputum samples were not graded.

**Table 2.** Secondary outcomes for comparison of tongue swab- vs. sputum-based molecular testing for TB (N=1639).

|  | **Tongue swab,**<br>**% (n/N)** | **Sputum<sup>a</sup>,**<br>**% (n/N)** | **Difference,**<br>**% (95% CI)** | **p-value** |
| --- | --- | --- | --- | --- |
| Provided sample | 99.9<br>(1637/1639) | 84.7<br>(1389/1639) | 15.1<br>(13.3, 16.9) | <0.001 |
| Valid molecular test result <sup>b</sup> | 98.3<br>(1611/1639) | 80.4<br>(1318/1639) | 17.9<br>(15.8, 20.0) | <0.001 |
| Experienced discomfort <sup>c</sup> | 8.6<br>(141/1638) | 21.8<sup>d</sup><br>(330/1511) | -13.2<sup>d</sup><br>(-15.8, -10.7) | <0.001 |

<sup>a</sup> Includes stool testing for young children (n=24)<sup>b</sup> MiniDock MTB for tongue swab and World Health Organization-recommended rapid diagnostic (Xpert MTB/RIF Ultra or Truenat MTB Plus) for sputum<sup>c</sup> n=1 did not complete preference survey<sup>d</sup> among n=1511 participants who attempted to produce sputum**Table 3. Sample collection preference overall and by subgroup (N=1638).**

The proportion of non-actionable molecular test results was similar for the tongue swab MiniDock MTB Test (26/1,637; 1·6%) and sputum WRD (27/1,389; 1·9%). However, due to differences in sample provision, the proportion of participants with a valid molecular test result overall was higher with tongue swabs (1,611/1,639; 98·3%) than with sputum (1,318/1,639; 80·4%; difference 17.9%, 95% CI 15·8-20·0) (**Table 2**).

Among those who attempted to provide both samples (n=1,511), a lower proportion reported discomfort providing tongue swabs (n=130; 8·6%) than providing sputum (n=330; 21·8%; difference -13·2%, 95% CI -15·8 to -10·7) (**Table 2**). Results were similar across subgroups (**Table S5**).

Most participants (1,022/1,638 or 62·4%) reported an overall preference for tongue swab collection, followed by no sample preference (314/1,638; 19·2%) and preference for sputum samples (302/1,638; 18.4%) (**Table 3**). The preference for tongue swab collection was observed across all subgroups. However, the proportion who preferred tongue swab collection varied particularly by country (range 45·9% in Vietnam to 82·3% in Uganda) and age group (range 46·5% in participants ≥65 years to 87·1% among caregivers of participants <5 years).

**Table 3.** Sample collection preference overall and by subgroup (N=1638).

|  | **Tongue swab** | **Sputum** | **No preference** | **p-value** |
| --- | --- | --- | --- | --- |
| Overall | 62.4% (1022/1638) | 18.4% (302/1638) | 19.2% (314/1638) |  |
| *Country* |  |  |  | <0.001 |
| Philippines | 64.1% (175/273) | 24.9% (68/273) | 11.0% (30/273) |  |
| Vietnam | 45.9% (298/649) | 20.5% (133/649) | 33.6% (218/649) |  |
| Uganda | 82.3% (247/300) | 17.0% (51/300) | 0.7% (2/300) |  |
| Zambia | 72.6% (302/416) | 12.0% (50/416) | 15.4% (64/416) |  |
| *Sex<sup>a</sup>* |  |  |  | 0.046 |
| Male | 60.0% (471/785) | 18.3% (144/785) | 21.7% (170/785) |  |
| Female | 64.6% (550/851) | 18.5% (157/851) | 16.9% (144/851) |  |
| *HIV Status* |  |  |  | <0.001 |
| Positive | 58.0% (240/414) | 17.6% (73/414) | 24.4 (101/414) |  |
| Negative | 68.6% (626/913) | 18.1% (165/913) | 13.4% (122/913) |  |
| Unknown | 50.2% (156/311) | 20.6% (64/311) | 29.3% (91/311) |  |
| *Age group* |  |  |  | <0.001 |
| <5 years | 87.1% (115/132) | 4.6% (6/132) | 8.3% (11/132) |  |
| 5-11 years | 81.3% (87/107) | 10.3% (11/107) | 8.4% (9/107) |  |
| 12-18 years | 76.7% (66/86) | 11.6% (10/86) | 11.6% (10/86) |  |
| 19-64 years | 58.8% (687/1169) | 21.1% (247/1169) | 20.1% (235/1169) |  |
| ≥65 years | 46.5% (67/144) | 19.4% (28/144) | 34.0% (49/144) |  |

<sup>a</sup> missing for n=2 who declined to report

### Sensitivity Analysis

We conducted a sensitivity analysis to assess diagnostic yield when altering our definition to include sputum Xpert results with a semiquantitative result of ‘Trace’ as positive. As expected, the diagnostic yield of sputum-based testing increased modestly from 4·1% to 4·9% (**Supplementary Table S6**). However, tongue swab-based molecular testing remained non-inferior to sputum-based testing (difference -1·0%; 95% CI -2·0, -0·06) overall and in most subgroups.

## DISCUSSION

In this pragmatic study at primary health centers in four high burden countries, we found that the diagnostic yield for TB of tongue swab-based testing using the MiniDock MTB Test was non-inferior to sputum-based testing with WRDs. Additionally, more people with presumptive TB were able to provide a tongue swab sample and had a valid molecular test result compared to routine sputum testing. These findings provide compelling evidence that tongue swab-based molecular testing is a viable alternative to sputum-based molecular testing for the diagnosis of pulmonary TB at the primary care level.

The primary outcome of our study demonstrated a comparable diagnostic yield between tongue swab- (3·8%) and sputum-based (4·1%) molecular testing, with a difference that fell within the pre-specified margin of non-inferiority. This finding is important because it suggests that despite the potentially lower sensitivity of tongue swab molecular tests reported in earlier studies, the higher rate of sample provision can offset this limitation, resulting in a similar overall diagnostic yield. This is particularly relevant in contexts where sputum sample collection is problematic, such as among young children, the elderly, and people living with HIV.^3,14^

Our study also highlights the substantial advantages of tongue swab-based testing in terms of sample provision and participant comfort. Notably, nearly all participants (99·9%) were able to provide a same-day tongue swab, whereas only 84·7% provided sputum. This difference underscores the practicality and accessibility of tongue swab collection, salient strengths given that the true impact of TB diagnosis on individuals and communities relies on the entirety of the diagnostic care cascade, not just test sensitivity.^15^ Moreover, participants reported less discomfort with tongue swabs compared to sputum collection, which may enhance compliance and reduce barriers to testing, particularly in children, people with minimal symptoms and for community-based screening programs.^16,17^

The implications of these findings are far-reaching for TB control programs, especially in high-burden countries where sputum smear microscopy remains widely used due to its low cost and minimal infrastructure requirements despite its lower sensitivity compared to molecular tests.^2^ Our data suggest that tongue swab-based molecular testing could, at a minimum, replace sputum smear microscopy, providing a more sensitive diagnostic tool that is also simpler and more acceptable to patients. This could lead to earlier and more accurate detection of TB cases, thereby improving treatment outcomes and reducing transmission.^18,19^

The MiniDock MTB Test can conceivably be implemented in settings with or without access to current sputum-based WRDs, because it is a relatively simple, portable, battery-operated platform, with rapid turnaround time of 12-25 minutes.^20^ The lower anticipated cost of the MiniDock MTB Test compared to current sputum-based WRDs could facilitate broader implementation and integration into existing TB diagnostic algorithms.^21^ Recently, it has been demonstrated that sputum swabs tested with the MiniDock MTB Test have similar sensitivity to sputum Xpert Ultra.^9^ Diagnostic yield is therefore likely to be even higher than what we report here with algorithms that prioritize testing of sputum when available.

Our study has several strengths, including its multi-country design, large sample size, and pragmatic approach, which enhance the generalizability and applicability of our findings. We also included diverse populations, encompassing children, men and women, and people living with and without HIV, which allowed for a comprehensive assessment of the diagnostic yield across different subgroups. However, there are limitations to consider. The reliance on a single novel tongue swab molecular assay (MiniDock MTB Test) means our results may not be directly generalizable to other swab-based platforms. Additionally, tongue swab sample collection and MiniDock MTB testing were done by research staff, and results could not be used to inform treatment decisions. Future studies are needed to verify our findings in the context of routine care and impact on person-centered outcomes including treatment initiation. Last, we did not perform reference standard testing to verify specificity. However, previous studies have consistently demonstrated high specificity of tongue swab-based molecular testing for TB,^6^ including with the MiniDock MTB Test.^9^

In conclusion, our study supports the adoption of tongue swab-based molecular testing as a feasible and effective alternative to sputum-based testing for TB diagnosis. The benefits of increased sample provision, reduced patient discomfort, and potential cost savings highlight the value of swab-based platforms, particularly in settings where sputum collection is difficult or where smear microscopy is still in use. These findings advocate for the integration of swab-based molecular tests into national TB programs to enhance diagnostic capacity and ultimately improve TB control efforts globally. Future research should focus on the long-term implementation outcomes and cost-effectiveness of swab-based TB diagnostics in various healthcare settings.

## Data Availability

All data produced in the present study are available upon reasonable request to the authors

## Acknowledgments

We gratefully acknowledge the individuals who participated in this study and also extend our sincere thanks to the administrative teams and clinical staff at the participating health centers for their support and dedication. This work would not have been possible without their partnership and collaboration.

## Data sharing

De-identified participant data that underlie the results reported in this article, study protocol, statistical analytical plan, and/or analytic code to researchers who provide a methodologically sound proposal. Proposals should be directed to; to gain access, data requestors will need to sign a data access agreement.

## SUPPLEMENTARY TABLES

**Table S1.** Diagnostic yield of tongue swab- vs. sputum based molecular testing for TB (N=1639).

|  | **Tongue swab MiniDock**<br>**MTB, % (n/N)** | **Sputum<sup>a</sup> WRD,**<br>**% (n/N)** | **Difference,**<br>**% (95% CI)** | **p-value** |
| --- | --- | --- | --- | --- |
| Overall | 3.8% (63/1639) | 4.1% (68/1639) | -0.3 (-1.2, +0.6) | 0.57 |
| *Country* |  |  |  |  |
| Philippines | 4.4% (12/273) | 5.1% (14/273) | -0.7 (-4.0, +2.5) | 0.80 |
| Vietnam | 2.5% (16/650) | 3.2% (21/650) | -0.8 (-2.0, +0.5) | 0.27 |
| Uganda | 1.0% (3/300) | 2.0% (6/300) | -1.0 (-2.5, +0.5) | 0.25 |
| Zambia | 7.7% (32/416) | 6.5% (27/416) | +1.2 (-1.0, +3.4) | 0.33 |
| *HIV status* |  |  |  |  |
| Positive | 2.9% (12/415) | 4.1% (17/415) | -1.2 (-3.0, +0.6) | 0.23 |
| Negative | 5.3% (48/913) | 4.8% (44/913) | +0.4 (-0.8, +1.7) | 0.58 |
| Unknown | 1.0% (3/311) | 2.3% (7/311) | -1.3 (-3.4, +0.8) | 0.29 |
| *Sex<sup>b</sup>* |  |  |  |  |
| Male | 5.0% (39/786) | 6.0% (47/786) | -1.0 (-2.5, +0.5) | 0.20 |
| Female | 2.8% (24/851) | 2.5% (21/851) | +0.4 (-0.8, +1.5) | 0.65 |
| *Age group* |  |  |  |  |
| <5 years | 0% (0/132) | 0% (0/132) | — | — |
| 5-11 years | 0.9% (1/107) | 0.9% (1/107) | 0.0 (-0.9, +0.9) | >0.99 |
| 12-18 years | 2.3% (2/86) | 2.3% (2/86) | 0.0 (-1.2, +1.2) | >0.99 |
| 19-64 years | 5.0% (58/1170) | 5.5% (64/1170) | -0.5 (-1.8, +0.7) | 0.47 |
| ≥65 years | 1.4% (2/144) | 0.7% (1/144) | +0.7 (-1.4, +2.7) | >0.99 |

<sup>a</sup>Includes stool testing for young children (n=24)
<sup>b</sup>Missing for n=2 who declined to report
Abbreviations: WRD, World Health Organization-recommended rapid diagnostic (Xpert MTB/RIF Ultra or Truenat MTB Plus; Xpert “trace” results considered negative)

**Table S2.** Secondary outcomes for comparison of tongue swab vs. sputum-based molecular testing for TB, by country (N=1639).

|  | Tongue swab,<br>% (n/N) | Sputum<sup>a</sup>,<br>% (n/N) | Difference,<br>% (95% CI) | p-value |
| --- | --- | --- | --- | --- |
| *Provided sample* |  |  |  |  |
| **Overall** | 99.9 (1637/1639) | 84.8 (1389/1639) | 15.1 (13.3, 16.9) | <0.001 |
| Philippines | 100 (273/273) | 64.5 (176/273) | 35.5 (29.5, 41.6) | <0.001 |
| Vietnam | 99.7 (648/650) | 97.7 (635/650) | 2.0 (0.6, 4.4) | 0.002 |
| Uganda | 100 (300/300) | 70.7 (212/300) | 29.3 (23.8, 34.8) | <0.001 |
| Zambia | 100 (416/416) | 88.0 (366/416) | 12.0 (8.7, 15.4) | <0.001 |
| *Valid molecular test result* |  |  |  |  |
| **Overall** | 98.3 (1611/1639) | 80.4 (1318/1639) | 17.9 (15.8, 20.0) | <0.001 |
| Philippines | 100 (273/273) | 64.1 (175/273) | 35.9 (29.8, 42.0) | <0.001 |
| Vietnam | 96.9 (630/650) | 97.4 (633/650) | -0.5 (-2.4, 1.5) | 0.74 |
| Uganda | 100 (300/300) | 70.7 (212/300) | 29.3 (23.8, 34.8) | <0.001 |
| Zambia | 98.1 (408/416) | 71.6 (298/416) | 26.4 (21.6, 31.3) | <0.001 |
| *Experienced discomfort<sup>c</sup>* |  |  |  |  |
| **Overall** | 8.6 (141/1638) | 21.8 (330/1511)<sup>d</sup> | -13.2 (-15.8, -10.7)<sup>d</sup> | <0.001 |
| Philippines | 4.4 (12/273) | 20.6 (42/204) | -19.1 (-25.5, -12.7) | <0.001 |
| Vietnam | 9.2 (60/650) | 17.2 (111/646) | -7.9 (-11.6, -4.2) | <0.001 |
| Uganda | 10.0 (30/300) | 27.4 (67/245) | -15.9 (-23.2, -8.6) | <0.001 |
| Zambia | 9.4 (39/416) | 26.4 (110/416) | -17.1 (-22.3, -11.8) | <0.001 |

<sup>a</sup> Includes stool testing for young children (n=24)<sup>b</sup> MiniDock MTB for tongue swab and World Health Organization-recommended rapid diagnostic (Xpert MTB/RIF Ultra or Truenat MTB Plus) for sputum<sup>c</sup> n=1 did not complete preference survey<sup>d</sup> among those who attempted to produce sputum (n=1511)

**Table S3.** Secondary outcomes for comparison of tongue swab- vs. sputum-based molecular testing for TB, by sex (N=1637)

|  | Tongue swab,<br>% (n/N) | Sputum<sup>a</sup>,<br>% (n/N) | Difference,<br>% (95% CI) | p-value |
| --- | --- | --- | --- | --- |
| *Provided sample* |  |  |  |  |
| **Overall** | 99.9 (1635/1637) | 84.9 (1389/1637) | 15.0 (13.2, 16.8) | <0.001 |
| Male | 99.9 (785/786) | 86.0 (676/786) | 13.9 (11.3, 16.4) | <0.001 |
| Female | 99.9 (850/851) | 83.8 (713/851) | 16.1 (13.5, 18.7) | <0.001 |
| *Valid molecular test result<sup>b</sup>* |  |  |  |  |
| **Overall** | 98.3 (1609/1637) | 80.5 (1318/1637) | 17.8 (15.7, 19.9) | <0.001 |
| Male | 98.1 (771/786) | 80.9 (636/786) | 17.2 (14.1, 20.3) | <0.001 |
| Female | 98.5 (838/851) | 80.1 (682/851) | 18.3 (15.4, 21.3) | <0.001 |
| *Experienced discomfort<sup>c</sup>* |  |  |  |  |
| **Overall** | 8.6 (141/1636) | 21.8 (329/1509)<sup>d</sup> | -13.2 (-15.7, -10.6)<sup>d</sup> | <0.001 |
| Male | 7.5 (59/785) | 19.1 (139/727) | -11.7 (-15.3, -8.1) | <0.001 |
| Female | 9.6 (82/851) | 24.3 (190/782) | -14.6(-18.3, -10.9) | <0.001 |

<sup>a</sup> Includes stool testing for young children (n=24)<sup>b</sup> MiniDock MTB for tongue swab and World Health Organization-recommended rapid diagnostic (Xpert MTB/RIF Ultra or Truenat MTB Plus) for sputum<sup>c</sup> n=1 did not complete preference survey<sup>d</sup> among those who attempted to produce sputum (n=1511)

**Table S4.** Secondary outcomes for comparison of tongue swab- vs. sputum-based molecular testing for TB, by HIV status (N=1639)

|  | Tongue swab,<br>% (n/N) | Sputum<sup>a</sup>,<br>% (n/N) | Difference,<br>% (95% CI) | p-value |
| --- | --- | --- | --- | --- |
| *Provided sample* |  |  |  |  |
| **Overall** | 99.9 (1637/1639) | 84.8 (1389/1639) | 15.1 (13.3, 16.9) | <0.001 |
| Positive | 100 (415/415) | 94.7 (393/415) | 5.3 (2.9, 7.7) | <0.001 |
| Negative | 99.9 (912/913) | 78.9 (720/913) | 21.0 (18.3, 23.8) | <0.001 |
| Unknown | 99.7 (310/311) | 88.8 (276/311) | 10.9 (7.0, 14.8) | <0.001 |
| *Valid molecular test result<sup>b</sup>* |  |  |  |  |
| **Overall** | 98.3 (1611/1639) | 80.4 (1318/1639) | 17.9 (15.8, 20.0) | <0.001 |
| Positive | 98.8 (410/415) | 88.9 (369/415) | 9.9 (6.4, 13.4) | <0.001 |
| Negative | 99.0 (904/913) | 74.3 (678/913) | 24.8 (21.7, 27.8) | <0.001 |
| Unknown | 95.5 (297/311) | 87.1 (271/311) | 8.4 (3.5, 13.2) | <0.001 |
| *Experienced discomfort<sup>c</sup>* |  |  |  |  |
| **Overall** | 8.6 (141/1638) | 21.8 (330/1511)<sup>d</sup> | -13.2 (-15.8, -10.7)<sup>d</sup> | <0.001 |
| Positive | 8.7 (36/414) | 20.9 (86/412) | -12.1 (-16.9, -7.4) | <0.001 |
| Negative | 7.5 (68/913) | 23.8 (193/812) | -15.8 (-19.3, -12.2) | <0.001 |
| Unknown | 11.9 (37/311) | 17.8(51/287) | -7.7 (-13.8, -1.5) | 0.014 |

<sup>a</sup> Includes stool testing for young children (n=24)<sup>b</sup> MiniDock MTB for tongue swab and World Health Organization-recommended rapid diagnostic (Xpert MTB/RIF Ultra or Truenat MTB Plus) for sputum<sup>c</sup> n=1 did not complete preference survey<sup>d</sup> among those who attempted to produce sputum (n=1511)

**Table S5.** Secondary outcomes for comparison of tongue swab vs. sputum-based molecular testing for TB, by age group (N=1639)

|  | **Tongue swab,**<br>**% (n/N)** | **Sputum<sup>a</sup>,**<br>**% (n/N)** | **Difference,**<br>**% (95% CI)** | **p-value** |
| --- | --- | --- | --- | --- |
| *Provided sample* |  |  |  |  |
| **Overall** | 99.9 (1637/1639) | 84.8 (1389/1639) | 15.1 (13.3, 16.9) | <0.001 |
| < 5 years | 100 (132/132) | 24.2 (32/132) | 75.8 (67.7, 83.8) | <0.001 |
| 5 – 11 years | 100 (107/107) | 66.4 (71/107) | 33.6 (23.8, 43.5) | <0.001 |
| 12 – 18 years | 100 (86/86) | 89.5 (77/86) | 10.5 (2.8, 18.1) | 0.004 |
| 19 – 64 years | 99.8 (1168/1170) | 92.1 (1078/1170) | 7.7 (6.0, 9.3) | <0.001 |
| ≥ 65 years | 100 (144/144) | 91.0 (131/144) | 9.0 (3.7, 14.4) | 0.002 |
| *Valid molecular test result<sup>b</sup>* |  |  |  |  |
| **Overall** | 98.3 (1611/1639) | 80.4 (1318/1639) | 17.9 (15.8, 20.0) | <0.001 |
| < 5 years | 100 (132/132) | 18.2 (24/132) | 81.8 (74.5, 89.2) | <0.001 |
| 5 – 11 years | 100 (107/107) | 56.1 (60/107) | 43.9 (33.6, 54.3) | <0.001 |
| 12 – 18 years | 98.8 (85/86) | 88.4 (76/86) | 10.5 (2.1, 18.9) | 0.012 |
| 19 – 64 years | 98.0 (1147/1170) | 88.0 (1029/1170) | 10.1 (7.9, 12.2) | <0.001 |
| ≥ 65 years | 97.2 (140/144) | 89.6 (129/144) | 7.6 (1.1, 14.1) | 0.019 |
| *Experienced discomfort<sup>c</sup>* |  |  |  |  |
| **Overall** | 8.6 (141/1638) | 21.8 (330/1511)<sup>d</sup> | -13.2(-15.8, -10.7)<sup>d</sup> | <0.001 |
| < 5 years | 12.1 (16/132) | 16.0 (8/50) | -2.0 (-19.2, +15.2) | >0.99 |
| 5 – 11 years | 4.7 (5/107) | 36.6 (30/82) | -32.9 (-45.9, -20.0) | <0.001 |
| 12 – 18 years | 2.3 (2/86) | 32.5 (27/83) | -30.1 (-41.7, -18.5) | <0.001 |
| 19 – 64 years | 8.8 (103/1169) | 20.5 (237/1154) | -11.6 (-14.5, -8.8) | <0.001 |
| ≥ 65 years | 10.4 (15/144) | 19.7 (28/142) | -9.2 (-17.9, -0.4) | 0.041 |

<sup>a</sup> Includes stool testing for young children (n=24)
<sup>b</sup> MiniDock MTB for tongue swab and World Health Organization-recommended rapid diagnostic (Xpert MTB/RIF Ultra or Truenat MTB Plus) for sputum
<sup>c</sup> n=1 did not complete preference survey
<sup>d</sup> among those who attempted to produce sputum (n=1511)

**Table S6.** Sensitivity analysis of diagnostic yield of tongue swab- vs. sputum-based molecular testing for TB (N=1639).

| **Diagnostic yield when Xpert MTB/RIF Ultra “trace”**<br>**results considered as positive** |  |  |  |  |
| --- | --- | --- | --- | --- |
|  | **Tongue swab MiniDock MTB,**<br>**% (n/N)** | **Sputum<sup>a</sup> WRD,**<br>**% (n/N)** | **Difference,**<br>**% (95% CI)** | **p-value** |
| Overall | 3.8% (63/1639) | 4.9% (80/1639) | -1.0 (-2.0, -0.06) | 0.036 |
| *Country* |  |  |  |  |
| Philippines | 4.4% (12/273) | 5.9% (16/273) | -1.5 (-4.9, +1.9) | 0.48 |
| Vietnam | 2.5% (16/650) | 4.3% (28/650) | -1.8 (-3.3, -0.4) | 0.008 |
| Uganda | 1.0% (3/300) | 2.0% (6/300) | -1.0 (-2.5, +0.5) | 0.25 |
| Zambia | 7.7% (32/416) | 7.2% (30/416) | +0.5 (-1.9, +2.8) | 0.82 |
| *HIV status* |  |  |  |  |
| Negative | 5.3% (48/913) | 5.5% (50/913) | -0.2 (-1.6, +1.2) | 0.87 |
| Positive | 2.9% (12/415) | 4.8% (20/415) | -1.9 (-3.9, +0.07) | 0.057 |
| Unknown | 1.0% (3/311) | 3.2% (10/311) | -2.3 (-4.4, -0.06) | 0.039 |
| *Sex<sup>b</sup>* |  |  |  |  |
| Male | 5.0% (39/786) | 7.0% (55/786) | -2.0 (-3.7, -0.4) | 0.014 |
| Female | 2.8% (24/851) | 2.9% (29/851) | -0.1 (-1.3, +1.1) | >0.99 |
| *Age group* |  |  |  |  |
| <5 years | 0% (0/132) | 0% (0/132) | — | — |
| 5 – 11 years | 0.9% (1/107) | 0.9% (1/107) | 0.0 (-0.9, +0.9) | >0.99 |
| 12 – 18 years | 2.3% (2/86) | 2.3% (2/86) | 0.0 (-1.2, +1.2) | >0.99 |
| 19 – 64 years | 5.0% (58/1170) | 6.3% (74/1170) | -1.4 (-2.7, -0.01) | 0.048 |
| ≥ 65 years | 1.4% (2/144) | 2.1% (3/144) | -0.7 (-2.7, +1.4) | >0.99 |

<sup>a</sup> Includes stool testing for young children (n=24)<sup>b</sup> missing for n=2 who declined to report
Abbreviations: WRD, World Health Organization-recommended rapid diagnostic (Xpert MTB/RIF Ultra or Truenat MTB Plus; Xpert “trace” results considered negative)

